# Individual attentional bias is positive or negative? It depends on trait anxiety

**DOI:** 10.1101/2025.04.24.25326135

**Authors:** Liping Hu, Menghui Xiong, Jianhui Liang, Yan Huang

**Author notes:** **Correspondence should be addressed to:** Yan Huang, 1068 Xueyuan Avenue, Nanshan District, Shenzhen, China, 518055.

## Abstract

The debate on whether the general population exhibits a positive or negative attentional bias has been ongoing. Some studies suggest that attentional bias is not necessarily a common characteristic among individuals, but rather varies on an individual basis. The aim of this study is to investigate the association between trait anxiety and attentional bias and its neural correlates. Seventy human adults with varying levels of trait anxiety participated in a novel emotional competition search task, where happy and angry faces were simultaneously presented amidst multiple neutral faces, with one emotional face as the target and another as the distractor. In the non-competition condition, one happy or angry face was presented among the neutral faces. An event-related potential (ERP) indicator of attentional selection, the N2pc, was used to measure individual attentional bias and to investigate whether this bias occurs in a top-down (towards targets) or bottom-up (towards distractors) manner. Our ERP results revealed a significant correlation between individuals’ levels of trait anxiety and their attentional biases. Specifically, lower levels of anxiety were associated with a stronger positive emotional bias, i.e., a larger N2pc for happy faces than angry faces, while higher levels of anxiety were linked to more pronounced negative bias. Additionally, this anxiety-associated attentional bias specifically manifested in bottom-up processes and in the emotion-competition condition. Our study provides electrophysiological evidence demonstrating that individual trait anxiety is associated with the emotional direction of attentional bias primarily through a stimulus-driven attentional process. This finding has implications for cognitive interventions targeting anxiety disorders.

## 1. Introduction

To deal with the overwhelming sensory information, our brains selectively prioritize specific information while disregarding the others. The emotional significance of sensory events is believed to exert a pivotal influence in the selection and inhibition of attentional resources (Vuilleumier, 2005). However, it remains debated whether the attention processing of the general population exhibits a bias towards positive or negative stimuli (Kauschke, Bahn, Vesker, & Schwarzer, 2019; Xu et al., 2021). Recent studies have investigated the influence of trait anxiety on negative bias by comparing threatening stimuli with neutral ones and found a correlation between trait anxiety and negative bias, with higher levels of anxiety linked to more pronounced negative biases (Veerapa et al., 2020; Wieser & Keil, 2020). A meta-analysis conducted by Bar-Haim et al. (2007) found that the combined effect size of the threat-related negative bias was significant in individuals with anxiety but not in nonanxious controls. Trait anxiety, a normally distributed trait, reflects a relatively stable and enduring tendency to experience anxiety (Spielberger, Gorsuch, Lushene, Vagg, & Jacobs, 1983; Vigneau & Cormier, 2008). These observations suggest that attention bias may represent more of an individual characteristic rather than a collective trait within the population. This individualization may partially explain the inconsistencies regarding whether the general population exhibits a positive or negative bias. Therefore, this study aimed to investigate whether individuals’ inclination towards positive or negative bias is associated with their level of trait anxiety and to elucidate the neural mechanisms underlying the individual attentional bias towards emotion by utilizing EEG. Specifically, the study aimed to identify the specific attentional processes in which the attentional bias occurs.

Attention is considered to contain top-down and bottom-up processes (Corbetta & Shulman, 2002; Theeuwes, 2010). Top-down attention refers to voluntary control based on current goals, such as attending to target stimuli, while bottom-up attention is involuntary in nature and driven primarily by the salience of stimuli, such as attending to prominent distractor stimuli. Previous electrophysiological studies have examined the mechanisms underlying attentional bias by investigating either the top-down or bottom-up attentional processes (Eimer & Kiss, 2007; Reutter, Hewig, Wieser, & Osinsky, 2017; Wieser, Hambach, & Weymar, 2018). The dot-probe task, in which an angry or happy face and a neutral face are simultaneously presented, followed by a dot as a probe target, has been widely utilized for studying the neural mechanism of attentional bias (Torrence & Troup, 2018). In this paradigm, emotional faces serve solely as distractors, allowing for the examination of only the bottom-up process of attention. Wieser, Hambach, & Weymar (2018) utilized the visual search paradigm to investigate attentional bias, focusing solely on the examination of emotional faces as targets, thus exclusively examining top-down attention processes without addressing bottom-up attention processes.

To date, there has been a scarcity of studies concurrently investigating both processes, leaving the question unanswered as to whether emotional bias is primarily associated with bottom-up stimulus capture or top-down attentional control. Therefore, our study employs EEG recordings to investigate the neural mechanisms underlying individual attentional bias by directly comparing top-down and bottom-up attentional responses to emotional stimuli.

To investigate the mechanism of attentional bias, our study employed a visual search paradigm to explore both top-down and bottom-up attention towards emotional faces, with emotional faces being presented as either targets or distractors. Desimone and Duncan (1995) proposed that increasing competition among multiple objects in a visual search task can enhance attentional biases. Therefore, we introduced a novel emotional competition search compared to the traditional non-competition search paradigm. In the traditional search, either a happy or an angry face is embedded among neutral faces (Barra, Coll, Barras, & Kerzel, 2017; Feldmann-Wustefeld, Schmidt-Daffy, & Schubo, 2011; Horstmann, & Becker, 2020). In the new competition condition, both a happy face and an angry face are simultaneously embedded among neutral faces to create an emotional competition pattern. We hypothesize that the emotional attentional bias will be more pronounced in the competition condition compared to the non-competition condition. Previous research has shown that the duration of stimulus presentation plays a crucial role in detecting attentional bias (Bar-Haim, Lamy, Pergamin, Bakermans-Kranenburg, & van IJzendoorn, 2007). For instance, Bantin, Stevens, Gerlach, & Hermann (2016) observed a decrease in bias towards threatening faces as the duration increased from short (< 200 ms) to long (> 1000 ms). This finding suggests that attentional bias mainly reflects early attentional processes, while longer presentation is influenced by later processes such as strategy and control processes. Therefore, in addition to manipulating emotional competition condition, we also employed a short stimulus duration (200 ms) to enhance the bias effect.

The N2pc, an ERP component originating from the posterior parietal cortex, is widely recognized as a reliable indicator of selective attention (Eimer, 1996; Luck & Hillyard, 1994). The N2pc component consistently demonstrates its superior sensitivity compared to behavioral measures when assessing attentional bias (Kappenman, Farrens, Luck, & Proudfit, 2014; Kappenman, MacNamara, & Proudfit, 2015; Reutter et al., 2017; 2019). For example, Kappenman et al. (2014) discovered that while reaction time did not display a bias towards threatening images, the N2pc demonstrated a significant shift of attention towards threat. In the present study, we quantified individual attentional bias by measuring the difference in N2pc amplitude between angry and happy faces. A larger N2pc amplitude for happy faces represents a positive bias, and vice versa for a negative bias. Moreover, the N2pc can also be used to study whether attentional bias occurs in top-down or bottom-up attentional processes (Reutter, Hewig, Wieser, & Osinsky, 2017; Wieser, Hambach, & Weymar, 2018). When emotional faces serve as targets, the N2pc evoked by emotional faces represents the goal-driven/ top-down attentional selection, and the N2pc difference between angry and happy faces reflects top-down attentional bias; conversely, when they act as distractors, it denotes stimulus-driven/ bottom-up attentional bias.

Overall, the aim of this study was to investigate the relationship between trait anxiety and individual attentional bias, along with its underlying neural mechanisms. We recruited 70 healthy participants with varying levels of trait anxiety. We employed a novel visual search paradigm incorporating emotional competition, where happy and angry faces were simultaneously presented. This was compared to the classical non-competition condition in which only one emotional face (either happy or angry) was embedded among several neutral faces. We hypothesized that emotional competition conditions would enhance the detection of positive and negative biases. By examining the association between anxiety levels and individual attentional bias in both top-down and bottom-up processes, we aim to elucidate whether trait anxiety is associated with the direction of attentional bias and to identify the specific attentional processes involved in this association.

## 2. Material and methods

### 2.1. Participants

Seventy healthy Chinese undergraduate students were recruited. They received a fixed compensation of 250 RMB for their time and participation in the research. All participants had normal or corrected-to-normal vision and were right-handed. Five subjects withdrew from the experiment, one citing personal discomfort, while the remaining four were excluded due to inadequate EEG data quality resulting from head movement and other factors. This resulted in a final sample size of 65 participants. The sample size was determined based on two previous studies investigating the correlation between the N2pc and trait anxiety, which employed sample sizes of 56 and 66, respectively (Hu et al., 2023; Kappenman, Geddert, Farrens, McDonald, & Hajcak, 2021). Taking into account potential exclusions, we opted for a sample size of 70 participants.

This study was conducted in accordance with the principles outlined in the World Medical Association Declaration of Helsinki and was approved by the Human Research Ethics Committee of the Shenzhen Institute of Advanced Technology (SIAT), Chinese Academy of Sciences. Prior to their participation, all participants were provided with detailed information about the experimental procedures and provided written informed consent.

### 2.2. Questionnaire

The levels of trait anxiety in the participants were assessed using the State-Trait Anxiety Inventory (STAI; Spielberger et al., 1983), a widely utilized self-evaluation questionnaire for measuring trait anxiety. Higher scores on the STAI indicate elevated levels of trait anxiety. The trait-anxiety scores of the participants ranged from 23 to 64, with a mean of 41 and a standard deviation of 10, which aligns with established norms for the general population (Spielberger et al., 1983). To ascertain the reliability of trait anxiety reports employing the STAI, Cronbach’s alpha was computed (Booth et al., 2017; Steinweg et al., 2021). Cronbach’s alpha for the STAI-T yielded a value of 0.901, indicating robust internal reliability of scale scores in this study.

### 2.3. Stimuli and Procedures

The facial stimuli used in the experiment were selected from the Karolinska Directed Emotional Faces database (Lundqvist, Flykt, & Öhman, 1998). The dataset consisted of 12 facial identities, with an equal distribution of 6 male and 6 female faces. Each identity had three different facial expressions: happy, angry, and neutral. To minimize any potential influence stemming from hair, neck, or background elements, all images were cropped at the hairline. Furthermore, to eliminate any confounding factors related to color perception variations among participants, grayscale versions of the stimuli were employed while ensuring that their average luminance remained consistent throughout.

Each participant completed two search tasks, with one task involving the search for happy faces and the other for angry faces. Both tasks were identical except for their different targets, and the order of these tasks was counter-balanced among participants. In each search display, six faces (equally distributed between male and female) were presented along a virtual circle with a radius of 6^°^. Two faces were positioned along the vertical centerline. There were two emotional competition conditions: competition condition (occurring in half of the trials) where a happy face, an angry face, and four neutral faces were simultaneously presented; non-competition condition where only one emotional face (either happy or angry, each occurring in a quarter of the trials) was randomly presented alongside five neutral faces. The positions of the six faces and emotional competition conditions were randomized across trials. Each task comprised 960 trials, resulting in a total of 1920 trials.

During the experimental procedure, the stimuli were presented on a 100-Hz monitor against a black background. Participants viewed the monitor from a distance of 80 cm. A gray fixation cross was consistently displayed at the center of the screen throughout the experiment. Each trial commenced with a central fixation point lasting for 500 ms, followed by an immediate presentation of a search display for 200 ms. Participants were instructed to maintain their gaze fixated on the central point and respond promptly by pressing either the left or right button to indicate whether or not they detected the target stimulus. They were given 2 seconds to provide their response after being presented with each search display. Reaction times (RTs) were recorded to assess participants’ response speed and efficiency.

### 2.4. EEG recording and analysis

EEG data were collected using a 64-channel system (Brain Products) with a sampling rate of 1000 Hz. The reference electrode was placed at FCz, and the impedance was maintained below 5 kΩ. Offline EEG processing and analysis were conducted utilizing MATLAB and the EEGLAB toolbox. The recorded EEG signals were resampled to 250 Hz and underwent band-pass filtering using a non-causal two-way least-squares FIR filter with a cutoff range of 0.1-40 Hz and a roll-off rate of 12 dB/octave. The signals were then re-referenced to the average of the left and right mastoid channels. To remove artifacts related to eye blinks, horizontal eye movements, and heartbeats, independent component analysis (ICA) was performed using the BINICA routine in EEGLAB. Components associated with these artifacts were identified and subsequently removed based on their spatial, spectral, and temporal characteristics. The analyzed epochs ranged from 200 ms before stimulus onset to 500 ms after stimulus onset, with a pre-stimulus window of 200 ms used for baseline correction. To further eliminate horizontal eye movements, trials with blinks and vertical eye movements exceeding ±70 µV at Fp1/Fp2 channels and horizontal eye movements exceeding ±30 µV at F9/F10 channels were excluded. Approximately 0.8% of trials were rejected based on these criteria. For the subsequent ERP analyses, there were over 100 valid trials for each condition.

The N2pc components were obtained by subtracting ipsilateral ERPs from contralateral ERPs with respect to the location of stimulus of interest (the target or the distractor). The N2pc evoked by the target was extracted from trials with a centerline distractor and lateral target, or no distractor and lateral target. Distractor-evoked ERPs were extracted from trials involving a centerline target and lateral distractor, or no target and lateral distractor. For measuring the N2pc, electrode sites P7, PO7, P8, and PO8 were selected as they exhibited the largest amplitudes for the component (see Fig. 1). In accordance with previous relevant studies on visual search (Hu et al., 2019, 2023; Gaspar & McDonald, 2014, 2018; Luo et al., 2019), the target- and distractor-evoked N2pc time window was separately selected based on the peak (i.e. negative extreme points of the difference waves during 150-450 ms) of the averaged difference waves across all participants and conditions, with a window duration of 40 ms centered around the peak latency. In our study, the time window for target-evoked N2pc was observed to be 315-355 ms, while that of distractor-evoked N2pc was found to be 300-340 ms. For each participant and condition, the amplitudes of the target-evoked and distractor-evoked N2pcs were computed as the mean values within the respective time windows. The peak latency of N2pc was defined as the time point corresponding to the peak of the difference waves for each participant under each condition within a time window of 150-450 ms. Additionally, to further minimize the possibility of a spurious positive finding (Gaspar & McDonald, 2018), we conducted analyses of signed areas measured in wider time windows (200-400 ms) subsequent to the peak-amplitude analyses (see supplementary materials).

**Fig. 1.**
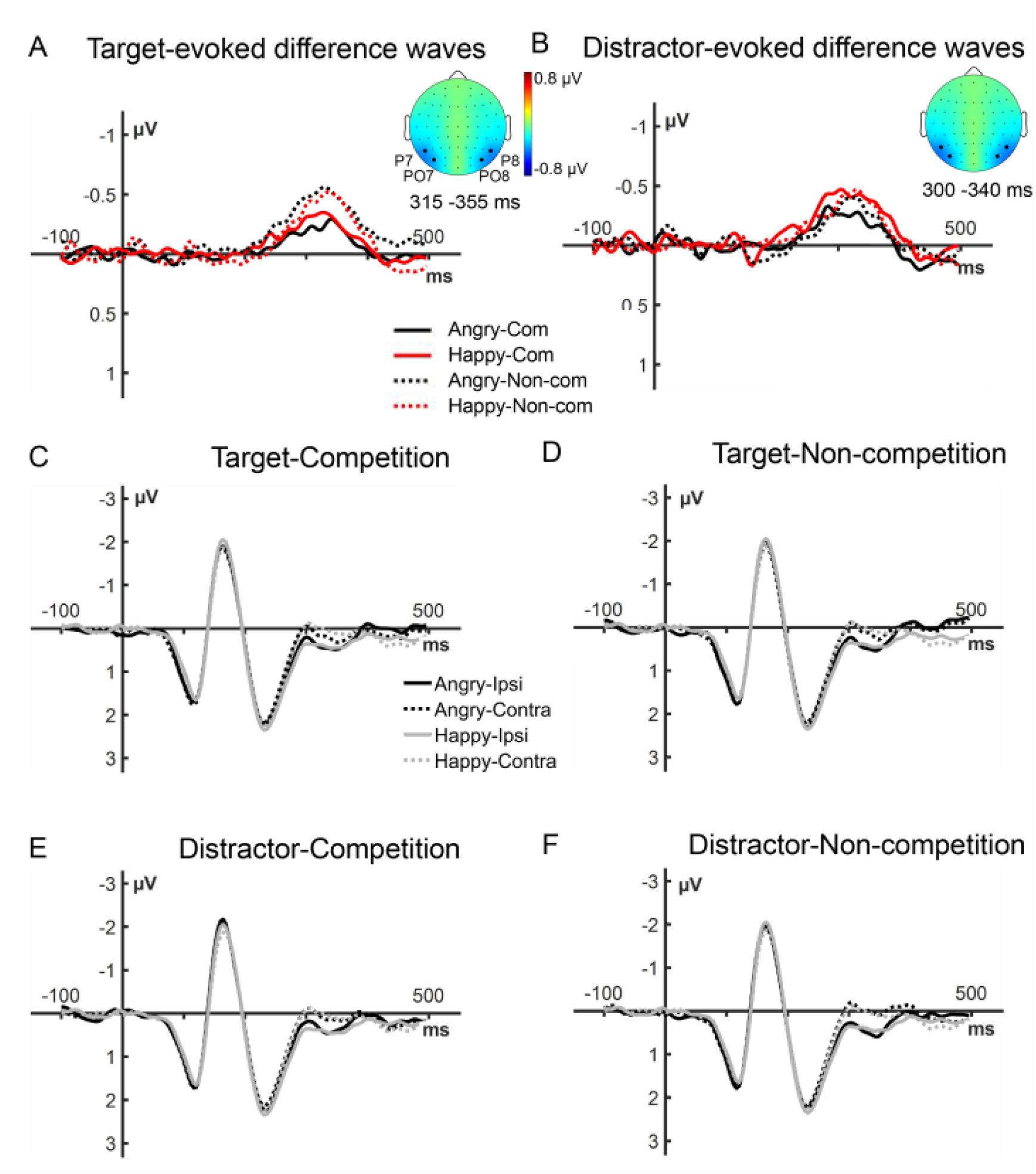
N2pc-based attentional bias and their correlation with trait anxiety levels (n = 65). ***A-B,*** The N2pc evoked by emotional faces as targets (**A**) and distractors (**B**). Contralateral-minus-ipsilateral difference waveforms of happy faces (red line) or angry faces (black line) are shown in the competition condition (solid line) and non-competition conditions (dotted line). The topographic maps illustrate the average N2pc across these four conditions within the specified time windows of the N2pc peaks. ***C-F,*** ERP waveforms recorded contralaterally and ipsilaterally to the angry or happy faces. We present the contralateral (dotted line) and ipsilateral (solid line) waveforms evoked by happy faces (gray line) or angry faces (black line) as targets in both the competition condition (**C**) and non-competition condition (**D**), as well as waveforms evoked by distractors in the competition condition (**E**) and non-competition condition (**F**).

### 2.5. Data Analysis

Data analysis mainly comprises two relatively independent parts: (1) Population level analysis. This part of the analysis was mainly to detect whether the variable competition affected attention to the targets or the distractors. Additionally, to observe whether there was an emotional bias of attention at the population level, so as to facilitate comparison with previous studies. We examined the effects of stimulus (target vs. distractor), competition (competition vs. non-competition), and emotion (happy vs. angry) on N2pc amplitudes and latency using a three-way repeated measures analysis of variance (ANOVA). Moreover, in order to test whether task order and individual anxiety level affected population-level attention, we further performed a four-way analysis of covariance (ANCOVA) incorporating task order (happy face search vs. angry face search) as a within-subject factor and anxiety levels as a continuous covariate. Detailed results of ANCOVA are provided in the supplementary materials. (2) Individual level analysis. To investigate how individual anxiety levels affect attentional bias, we conducted Pearson’s correlation analyses to explore potential associations between anxiety scores and ERP measures. P-values were adjusted using the false discovery rate (FDR) method at a threshold of 0.05 (Benjamini & Hochberg, 1995) to control for Type I errors. Additionally, Fisher’s r-to-z transformation (Lee & Preacher, 2013; Steiger, 1980) was employed to compare correlation coefficients using the online calculator available at https://quantpsy.org/corrtest/corrtest2.

To further examine the consistency of the various measurements within an individual, we also assessed the internal consistency reliability of behavioral and ERP measures with a split-half approach, where the correlation between averages of odd- and even-numbered trials was determined and corrected using the Spearman-Brown prophecy formula (Kappenman et al., 2021; Nunnally et al., 1967).

### 2.6. Transparency and openness

We report how we determined our sample size, all data exclusions, all manipulations, and all measures in the study. All data are available on OSF at https://osf.io/38p6a/. This study was not preregistered.

## 3. Results

### 3.1. ERP results

#### 3.1.1. Population level analysis

We measured attention on the target or the distractor by the peak amplitude and latency of the N2pc component. As depicted in Fig. 1A and B, both happy and angry faces, whether presented as targets or distractors, elicited a significant N2pc component under both competition and non-competition conditions (one sample t-test, *p*s < 0.001). To examine the influence of stimulus (target vs. distractor), competition (competition vs. non-competition), and emotion (happy vs. angry) on mean N2pc induced in the general population, a three-way ANOVA was conducted on mean peak amplitudes and latency, respectively. The results of peak amplitudes revealed that the main effect of competition was significant (F(1, 62) = 14.818, *p* < 0.001, η² = 0.188), showing a smaller amplitude in the competition condition as compared with the non-competition condition. Additionally, the interaction between competition and stimulus was significant (F(1, 62) = 7.233, *p* = 0.009, η² = 0.102), indicating that the impact of competition on the attention devoted to the target and the distractor varies. The main effect of emotion was not significant (F(1, 62) = 3.351, *p* = 0.072, η² = 0.05); however, there was a positive bias tendency, indicating that happy faces elicited larger peak amplitudes compared to angry faces, suggesting a weak positive bias at the population level. Also, emotion exhibits no significant interaction with other variables (*p*s > 0.1), suggesting that competition and stimulus did not affect directly the population-level attentional bias. Nevertheless, the results of the ANCOVA with anxiety level as a covariate reveal a significant four-way interaction among anxiety level, stimulus, competition and emotion (*p* = 0.039), suggesting that anxiety level exerts a significant influence on the effect of these factors on attention. In sum, the above results suggest that at the population level, there was no robust attentional bias, and competition significantly regulated attention, and its effect on the attention of the target and the distractor varied.

In addition to the peak amplitude approach, we also employed the signed area method with a broader time window (200-400 ms) to calculate the size of N2pc. This area-based amplitude is considered to be capable of accommodating greater inter-participant variability in the timing of ERP components (Luck, 2014). The results of the area amplitude analyses were consistent with those of the peak amplitude analyses (see supplementary materials).

The peak latency of the N2pc elicited by target or distractor faces was analyzed. The same three-way ANOVA was performed with the predictors of stimuli, competition, and emotion. The results revealed that the main effects of three factors and their interactions were not significant (*p*s ≥ 0.149), suggesting that these factors had no impact on N2pc latency.

#### 3.1.2. Individual level analysis

The primary objective of this study is to investigate whether positive or negative bias is associated with the level of individual trait anxiety. Hence, we further conduct correlation analyses between individual attentional bias and their trait anxiety level under the four conditions (competition × stimulus), respectively. The attention bias was computed by subtracting the N2pc amplitude elicited by happy faces from that induced by angry faces. Top-down and bottom-up attentional processes were respectively explored through target- and distractor-evoked N2pc. The results revealed that regarding the target-evoked N2pc, no significant correlations existed between attentional bias and anxiety level, both in the competition (*r* = −0.056, *p* = 0.660, Fig. 2A) and non-competition conditions (*r* = −0.077, *p* = 0.541, Fig. 2B). These findings suggest that top-down attentional bias did not vary with trait anxiety level. However, regarding the distractor-evoked N2pc (Fig. 2C and 2D), a significant correlation between attentional bias and trait anxiety level in the competition condition was observed (*r* = −0.392, *p* = 0.001, FDR-corrected *p* = 0.004), whereas no significant correlation was found in the non-competition condition (*r* = 0.001, *p* = 0.992). We further conducted statistical analyses to compare the correlation coefficients. For the distractor-evoked N2pc, the correlation between anxiety level and attentional bias was significantly more pronounced in the competition condition compared to the non-competition condition (z = −2.33, *p* = 0.020). Moreover, under the competition condition, the correlation coefficient for the distractor-evoked N2pc was significantly stronger than that for the target-evoked N2pc (z = −2.16, *p* = 0.030). For the area amplitudes of N2pc, we obtained consistent results, demonstrating a significant correlation between anxiety level and distractor-evoked N2pc (*r* = −0.316, *p* = 0.010), while no other significant correlations were identified (see supplementary materials). These findings imply that the attention bias of individuals is associated with their level of trait anxiety, and this correlation predominantly emerges in the bottom-up attention process. Moreover, the competition between positive and negative emotional faces is beneficial for the observation of this correlation.

**Fig. 2.**
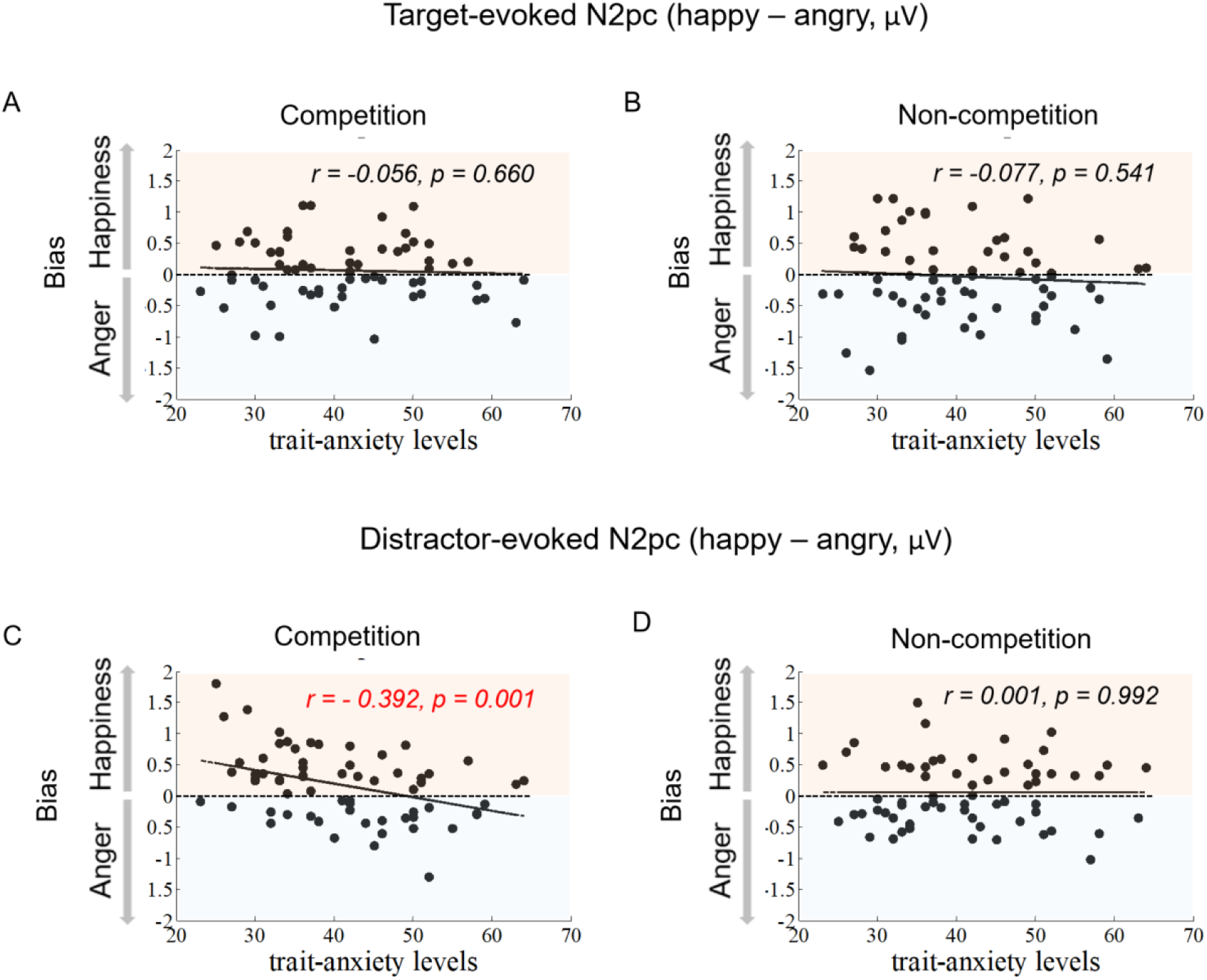
Attentional bias was quantified by calculating the difference in the peak amplitude of the N2pc elicited by angry and happy faces. When emotional faces served as targets, no significant correlations were found between attentional bias and trait-anxiety levels under both competition (**A**) and non-competition conditions (**B**). Nevertheless, when emotional faces acted as distractors, a significant correlation emerged between attentional bias and trait-anxiety levels specifically in the competition condition (**C**), while no significant correlation was observed in the non-competition condition (**D**).

Correlation analysis was also carried out on the peak latency of target-evoked and distractor-evoked N2pc to examine the association between individual attentional bias and their trait anxiety level. The results indicated no significant correlation either for target- or distractor-evoked N2pc under competition or non-competition conditions (*p*s ≥ 0.124).

### 3.2. Behavior results

The average hit rate for the target ranged from 65% to 70%, which aligns with the level of acuracy observed in previous visual search studies involving a presentation duration of 200 ms for faces (Feldmann-Wustefeld et al., 2011). A two-factor ANOVA was conducted to examine the effects of emotion (happy face search vs. angry face search) and competition (competition vs. non-competition). The results revealed a significant main effect of competition (competition vs. non-competition: 66.2% ± 1.6% vs. 69.5% ± 1.6%, *F* (2, 64) = 41.892, *p* < 0.001, η_P_ ^2^ = 0.396), while the main effect of emotional and their interaction were not significant (*p*s ≥ 0.073), indicating an interference effect from distractors in the competition condition. These findings suggest the absence of top-down attentional bias in behavior, which aligns with our electrophysiological results.

Similarly, the RT data were subjected to a 2-way ANOVA. Only the main effect of competition was found to be significant (competition vs. non-competition: 578 ±24 ms vs. 570 ± 23 ms, *F*(2, 64) = 5.747, *p* = 0.019, η_P_^2^ = 0.082), while the main effect of emotion and the interaction were not significant (*p*s ≥ 0.246). Both accuracy and RT results indicate the absence of a top-down attentional bias, which aligns with our EEG findings.

### 3.3. Internal consistency reliability of behavioral and ERP measures

We initially assessed the internal consistency reliability of behavioral and ERP measures. The internal consistency reliability of behavioral and ERP measures was evaluated using a split-half approach, wherein we determined the correlation between averages of odd- and even-numbered trials and subsequently corrected it utilizing the Spearman-Brown prophecy formula (Kappenman, Geddert, Farrens, McDonald, & Hajcak, 2021; Nunnally, 1967). The Spearman-Brown corrected correlations for behavioral and ERP measures were all statistically significant (all *r* > 0.837, all *ps* < 0.001). These results indicate that both behavioral and ERP measures in this study exhibit robust internal reliability.

## 4. Discussion

The aim of this study was to investigate the association of trait anxiety between individual attentional bias and its neural correlates. Employing an ERP indicator of selective attention, our findings revealed a significant association between individuals’ attentional bias and their levels of trait anxiety. Specifically, individuals with lower anxiety levels exhibited a stronger positive bias, whereas those with higher anxiety levels demonstrated a more pronounced negative attentional bias. These results suggest that attentional bias is individualized and is associated with varying levels of trait anxiety. Furthermore, we observed that the correlation between trait anxiety and attentional bias is predominantly evident in the bottom-up attentional process rather than the top-down attentional process. Notably, it is worth highlighting that the trait anxiety-associated attentional bias was solely evident in conditions involving emotional competition but disappeared when emotional faces were presented alone.

As discussed in recent reviews by Kauschke et al. (2019) and Xu et al. (2021), there is an ongoing debate on whether the general population exhibits a positive or negative bias. Previous studies have reported that trait anxiety affects the extent of negative bias, as evidenced by Dennis & Chen (2009) and Berggren & Eimer (2021). In contrast, in our current investigation, the correlation analysis revealed positive bias in individual with lower anxiety and negative bias in individual with higher anxiety. Although there was a tendency towards positive bias throughout the entire population in our study (*p* = 0.072), we contend that significant individual variations in attentional bias undermine the stability of any overall population-level attentional bias. This suggests that discussions about attentional bias in the general population may be limited, as the direction of attentional bias are associated with individual trait anxiety levels. In other words, attentional bias may not be a ubiquitous characteristic of the population, but rather an individual trait.

Previous studies have identified a relationship between anxiety and negative bias towards threat (Salahub & Emrich, 2020; Veerapa et al., 2020); however, our research extends beyond these findings. By utilizing both negative and positive face stimuli, we discovered that as trait anxiety levels increased among participants, there was a noticeable shift from a positive attentional bias to a negative one. To investigate whether this change in attentional bias was driven by attention to happy or angry faces alone, we conducted a correlation analysis between trait anxiety and N2pc responses elicited by happy or angry faces, respectively (see Table S1). We observed a correlated tendency between trait anxiety and N2pc amplitude for happy face distractors (r = 0.242, *p =* 0.052) and for angry face distractors (r = −0.234, *p =* 0.061) in the competitive condition. When compared to the stronger correlation between anxiety and the N2pc difference between happy and angry faces, these findings suggest that anxiety-related attentional bias is not solely influenced by one type of emotional stimulus. Instead, it reflects a dynamic interplay between attention directed toward both positive and negative stimuli, which varies with levels of trait anxiety. In summary, anxiety-related attentional biases arise from a shift in the balance of attention toward positive and negative stimuli. Our findings offer fresh insights into the neural mechanisms underlying attentional bias in individuals with differing levels of trait anxiety.

In our study, we observed the effect of trait anxiety on attentional bias when emotional faces served as distractors rather than targets, suggesting that this effect predominantly manifests in the bottom-up attentional process rather than the top-down process. Previous investigations have primarily focused on a single attentional process. For instance, some prior research exploring the bottom-up process has reported attentional bias towards negative stimuli (Berggren & Eimer, 2021; Reutter et al., 2017). Conversely, other studies investigating the top-down process have found attentional bias towards general emotional stimuli without specific emphasis on negative or positive stimuli (Wieser, Hambach, & Weymar, 2018). In contrast, our study directly compares the top-down and bottom-up attentional processes within the same experimental context, thereby providing a distinct advantage in elucidating the predominant mechanism underlying attentional bias. By concurrently observing these two attentional processes, we reveal that the primary source of attentional bias originates from the bottom-up attention process. Similarly, a behavioral study on visual search examined the attentional bias towards emotional faces as both task-related and task-independent stimuli. The results indicated that trait anxiety primarily influenced the attentional bias in task-independent situations, highlighting its influence on bottom-up attentional process (Dodd, Vogt, Turkileri, & Notebaert, 2017). Essentially, these findings suggest that stimulus-driven (“bottom-up”) processes may be more indicative of anxiety traits than goal-oriented (“top-down”) processes.

It was observed that the association between trait anxiety and attentional bias exclusively manifested itself in the condition of positive and negative emotional competition, while it disappeared in a non-competitive setting. This finding suggests the emotional competition is more effective in eliciting attentional bias, which aligns with previous behavioral observations (e.g., Pinkham et al., 2010). Previous EEG studies examining attentional bias have typically employed non-competitive paradigms in which angry or happy emotional faces were presented alongside neutral faces (Brosch, Pourtois, Sander, & Vuilleumier, 2011, Liu et al., 2021). While these studies did observe a heightened N2pc response to emotional faces compared to neutral faces, no discernible distinction was detected between the effects of positive and negative emotions. Our electrophysiological investigation provides neural evidence that emotional competition elicits a heightened sensitivity towards attentional bias, thereby offering valuable insights for future research on this topic. Although non-competition trials were randomly interspersed with competition trials, in order to eliminate any potential inter-trial priming effects from exposure to a single emotional face in the non-competition trials, it is recommended that future research focus exclusively on competition conditions for a better exploration of emotional bias.

It is important to consider that the investigation of attentional bias can be influenced by a range of experimental stimuli, presentation conditions, and types of anxiety. For instance, Burra et al. (2016, 2017) utilized the visual search paradigm in their investigation of attentional bias; however, they observed that attentional bias was linked to distinct ERP components. Specifically, cartoon faces elicited N2pc components, while naturalistic faces evoked Pd components. Consequently, within the similar paradigm but with different stimulus types, they identified associations between N2pc and Pd and negative bias, respectively. Moreover, the duration of stimulus presentation may also induce different ERP components. In contrast to the findings of Burra et al. (2017), which reported that long-term exposure to emotional faces induced Pd, our current study observed that brief presentation of emotional faces for 200 ms elicited N2pc but not Pd. Additionally, different types of anxiety may have distinct influences on attentional bias. For example, in the dot-probe task using facial stimuli, it was observed that attentional bias towards threatening stimuli exhibited a correlation not with trait anxiety scores, but rather with levels of social anxiety (Kappenman et al., 2014; Reutter et al., 2017). Subsequent studies could further integrate diverse forms of anxiety and stimuli in order to elucidate the breadth of the present findings.

In terms of the association between trait anxiety and the bottom-up, stimulus-driven attentional capture, we propose a hypothesis that trait anxiety influences this process through selection history. Our attention is more likely to be drawn towards objects that have been previously attended, which is the effect of selection history on attention. Previous studies have suggested that the selection history exerts an influence on attention through its impact on current objectives and the salience of stimuli (for a review, see Awh, Belopolsky & Theeuwes, 2012; Failing & Theeuwes, 2018). Notably, both positive and negative emotional stimuli possess similar levels of salience in terms of their physical characteristics; therefore, they should elicit comparable degrees of attention capture without introducing any bias in the stimulus itself. However, the selection history of individuals with varying levels of trait anxiety diverges. Individuals exhibiting high trait anxiety demonstrate a heightened frequency of negative emotions in their choice history, whereas those with low anxiety exhibit a greater occurrence of positive emotions in their decision-making process (Vigneau & Cormier, 2008). This disparity in selection history results in stimuli with comparable physical salience evoking distinct degrees of salience, thereby eliciting differential attention capture. That is, individuals with high anxiety have a positive bias while individuals with low anxiety have a negative bias. Furthermore, this attentional bias perpetuates and reinforces the individual’s selection history. In the complex interplay between selection history and attention processes, attentional bias is further strengthened.

Repeated reinforcement of negative bias is widely acknowledged as a crucial factor in the development and maintenance of anxiety disorders (Bar-Haim et al., 2007; Hakamata et al., 2010). The extensively discussed attention bias modification (ABM) training aims to alleviate anxiety by attenuating negative bias (MacLeod, Rutherford, Campbell, Ebsworthy, & Holker, 2002; Macleod & Clarke, 2015; Mogg & Bradley, 2018). This strategy involves instructing individuals to actively ignore threatening stimuli through top-down attentional control. Here, we propose that intentionally increasing the selection history of positive stimuli could potentially serve as an effective way to alleviate anxiety. Building upon our recent findings that individuals with low levels of anxiety exhibit a positive attentional bias, we propose the implementation of positive bias training (PBT) as a means to reverse negative bias. In PBT, we aim to counteract the negative bias in individuals with high anxiety by increasing their exposure to attentional capture scenarios involving positive stimuli, thereby augmenting the frequency of positive stimuli in their selection history. This approach has the potential to modify an individual’s negative bias and may offer assistance in mitigating anxiety.

In conclusion, our findings provide valuable electrophysiological evidence that underscores the role of individual trait anxiety level in shaping attentional bias and elucidate the predominant mechanism underlying such biases. Future research could investigate the potential impact of this enduring negative attentional bias on the occurrence and progression of emotion-related disorders, as well as explore cognitive training interventions aimed at ameliorating the negative bias.

## Acknowledgments

This study was supported by STI2030-Major Projects (2022ZD0209500), National Natural Science Foundation of China (32371091), Basic and Applied Basic Research Foundation of Guangdong Province (2024A1515010529, 2023A1515012642), STI2030-Major Projects (2021ZD0200700), Guangdong Provincial Key Laboratory of Brain Connectome and Behavior (2023B1212060055), and CAS Key Laboratory of Brain Connectome and Manipulation (2019DP173024).

## Author contributions

**Liping Hu** and **Yan Huang** designed the study, analyzed the data, and wrote the paper. **Menghui Xiong** and **Jianhui Liang** conducted the experiments. All authors contribute to and have approved the final text.

## Data availability

All data are available on OSF at https://osf.io/38p6a/.

## Supplementary Materials

### 1. ANCOVA on the amplitudes and latency of the N2pc

#### N2pc Amplitude

To test whether task order and individual anxiety level had an impact on population-level attention, we conducted a four-way ANCOVA, with factors including stimulus (target vs. distractor), competition (competition vs. non-competition), emotion (happy vs. angry), and task order (happy face search vs. angry face search), incorporating anxiety levels as a continuous covariate. The results indicated that the main effect of task order was not significant (*p* = 0.452), and no any significant interactions involving task order were observed (*p* ≥ 0.097). These findings suggest that task order had no effect on attention as indexed by the N2pc amplitude. Additionally, a significant four-way interaction among anxiety level, stimulus, competition and emotion was detected (*p* = 0.039), suggesting that trait anxiety level had an impact on the effect of these factors on attention.

#### N2pc Peak Latency

The results of the four-way ANCOVA on N2pc peak latency indicated that the main effect of task order and its interactions with the other predictors were non-significant (*p* ≥ 0.1), suggesting that task order exerted no significant influence on N2pc peak latency. Moreover, there was no significant effect related to anxiety level *(p* ≥ 0.2). These findings suggest that these experimental conditions have no significant impact on N2pc peak latency.

### 2. ANOVA and correlation analyses based on the area amplitudes of N2pc

Compared to the conventional mean-amplitude approach, the signed area method accommodates greater inter-participant variability in the timing of ERP components (Luck, 2014). For each participant, we quantified the N2pc as the negative area of the waveform that falls below the zero line within the 200-400 ms time window, which encompasses the conventional measurement windows. The results of three-way ANOVA revealed that the main effect of competition and the interaction between competition and stimulus were significant (*p* ≤ 0.001). The main effect of emotion was significant (F(1, 62) = 5.096, *p* = 0.027), indicating a positive bias at the population level, which is in line with the positive bias trend of the peak amplitude results. The other interactions were not significant (*p*s > 0.05). We also conducted correlation analyses between the N2pc area amplitudes and individual trait levels under the four conditions (competition ×stimulus). The results indicated that only the distractor-evoked N2pc exhibited a significant correlation with individual anxiety levels under the competition condition (*r* = −0.316, *p* = 0.01), whereas the correlations between the N2pc and anxiety level under the other three conditions were not significant (*p*s > 0.2). In sum, the results of the area amplitude analyses were consistent with those of the peak amplitude analyses.

### 3. The respective correlations of the N2pc elicited by happy or angry faces and the level of trait anxiety

The correlations between trait anxiety and the mean amplitude of N2pc elicited by happy or angry faces as distractors or targets in competition or non-competition conditions are detailed in Table S1. Our findings indicate that in the competition condition, both N2pcs induced by happy faces and angry faces as distractors were marginally correlated with anxiety levels (happy: r = 0.242, *p* = 0.052; angry: r = −0.234, *p* = 0.061). No other marginally significant or significant correlations were observed in the remaining conditions.

---

**Table S1.** Correlations between trait anxiety and N2pc evoked by happy or angry faces.

| Mean<br>amplitude | happy face |  |  |  | angry face |  |  |  |
| --- | --- | --- | --- | --- | --- | --- | --- | --- |
|  | target |  | distractor |  | target |  | distractor |  |
|  | com | non | com | non | com | non | com | non |
|  | -.044 | .080 | **.242** | .028 | -.121 | -.005 | **-.234** | .034 |
|  | .727 | .528 | **.052** | .822 | .335 | .967 | **.061** | .790 |

Note: ‘com’ represents competition condition, while ‘non’ corresponds to non-competition condition. The values in the first row represent the correlation coefficients, while the values in the second row correspond to the p-values.

